# The Current State of Timely Results Reporting Among Hypertension Trials on ClinicalTrials.gov: A Cross-Sectional Meta-Research Analysis

**DOI:** 10.64898/2026.08.05.26359829

**Authors:** William Harris, Parker Bragg, Lauren Kocour, Tanner Livsey, Ryan Langerman, Noah Calvert, Makalie Lackey, Amy Nguyen, Alicia Ito Ford, Matt Vassar

## Abstract

**Objectives:** To characterize how completely and promptly summary results are reported for registered hypertension trials on ClinicalTrials.gov, and whether reporting correlates with the observable obligation to report.

**Methods:** Cross-sectional analysis of completed or terminated interventional trials for hypertension, retrieved through the ClinicalTrials.gov API version 2. Trials required a primary completion date of type ACTUAL at least 12 months before extraction. Reporting was timed from primary completion to first results submission and classified as timely at 365 days or fewer. Applicability was approximated requiring interventional design, phase 2 or later, a United States site, and an FDA-regulated drug or device, assigned flag-confirmed or inferred. Proportions are reported with Wilson 95% confidence intervals, time to reporting by Kaplan-Meier, and adjusted associations by logistic regression clustered on lead sponsor.

**Results:** Of 5,851 trials, 5,396 were due to report. Timely reporting was 9.1% (95% CI 8.3-9.9) and any-time reporting 28.8% (95% CI 27.6-30.0). Reporting was graded by applicability, with flag-confirmed trials reporting timely at 36.9% (95% CI 31.6-42.5) and non-applicable trials at 6.3% (95% CI 5.6-7.1). A United States site carried the strongest adjusted association with timely reporting (OR 4.03, 95% CI 2.99-5.42). Among unreported trials, 7.8% had a sponsor-tagged publication and 36.4% under a broader definition.

**Conclusion:** Prompt registry reporting of hypertension trial results remains uncommon, and reporting is most closely associated with the observable obligation to report.

## Introduction

Hypertension is the most prevalent modifiable risk factor for cardiovascular disease and premature death, affecting an estimated 1.4 billion adults worldwide.^1,2^ Nearly every management decision, from which drug to initiate to what blood-pressure target to pursue, rests on evidence from clinical trials. When results are withheld or delayed the visible evidence skews toward favorable findings because trials with unfavorable or null results are the least likely to be reported.^3,4^ For a condition as common as hypertension, even modest distortion of the trial record propagates into guideline recommendations and prescribing decisions at scale.

The Food and Drug Administration Amendments Act of 2007 (FDAAA) was enacted to prevent this distortion, requiring the responsible party of an applicable clinical trial to submit summary results to ClinicalTrials.gov within twelve months after primary completion, an obligation clarified and extended by the 2017 Final Rule (42 CFR Part 11).^5–7^ Unlike journal publication, registry reporting presents results in a structured, searchable form on a fixed clock whether or not a manuscript is accepted, so a completed trial reaches the public record regardless.

National audits of this requirement have found reporting falls well short of the standard, with compliance concentrated among applicable clinical trials.^8^ Estimates vary, however, depending on how the obligated population is defined and whether timeliness is measured from results submission or results posting.^9,10^ These audits characterize the registry as a whole. Condition-level audits have begun to appear in other clinical areas but few, if any, have been conducted for hypertension.^11^

We conducted a cross-sectional meta-research study of results reporting among completed and terminated interventional hypertension studies registered on ClinicalTrials.gov. We described reporting across several criteria, estimated adjusted associations with timely reporting, and assessed reporting rates against published national reference values.

## Methods

### Study Design

This cross-sectional analysis evaluated the reporting of summary results for hypertension clinical trials registered on ClinicalTrials.gov. The OSU-CHS Institutional Review Board (#2026065) concluded that it did not qualify as research involving human subjects; as a result, informed consent was not required, and it was registered on the Open Science Framework (OSF). The frozen dataset, analysis code, and derived outputs are archived on OSF and GitHub at methodology-c c81585d/hypertension-frozen-v3.^12,13^ Detailed information on the methodology of this study can be found in Supplementary File 1.

### Data Source and Search Strategy

ClinicalTrials.gov API version 2 was used to retrieve trial records programmatically.^14^ Eligible records were identified using the registry condition query “hypertension,” which was preserved verbatim to allow exact reproduction of the cohort. As ClinicalTrials.gov maps condition searches through concept and synonym expansion rather than exact string matching, the cohort represents all records indexed by the registry to hypertension. Following extraction, the dataset was frozen and archived with complete provenance metadata, including the query string, API endpoint, execution timestamp (UTC), software version, and content hash.

### Eligibility and Reporting Denominator

Eligible studies were interventional trials with an overall status of completed or terminated. Terminated trials were retained but analyzed separately from completed trials. Inclusion in the reporting denominator further required an ACTUAL primary completion date that preceded data extraction by at least 12 months, thereby ensuring that each trial had reached the applicable reporting window. Records without an ACTUAL primary completion date were included in descriptive cohort characteristics but excluded from analyses requiring reporting-time calculations, and all exclusions were recorded.

### Data Extraction

All study variables were derived directly from structured ClinicalTrials.gov data fields, eliminating the need for manual data abstraction. The extracted dataset included trial status, study phase, primary completion date and date type, results submission and posting dates, registry indicators of results availability, lead sponsor information, FDA-regulated device and drug indicators, enrollment, intervention type, study locations, and linked references. Registry dates recorded at partial precision were assigned to the midpoint of the period by a fixed rule, and every imputation was tallied. Record counts, processing steps, and analytic decisions were documented throughout the workflow and archived with the frozen dataset.

### Variable Definitions

Trials were considered reported if the registry indicated results availability or a results-first-posted date. Reporting time was defined as the interval from ACTUAL primary completion to first results submission, with timely reporting defined as submission within 365 days; posting-based timing was evaluated in a sensitivity analysis. Sponsor type was classified as industry or non-industry using the lead sponsor field, refined with a standardized sponsor dictionary. Enrollment was analyzed on the base-2 logarithmic scale, excluding records with missing or zero values from the regression only. Linked publications were identified using both a strict definition (sponsor-tagged results references) and a broader definition that additionally included auto-derived PubMed links.

### Identification of Applicable Clinical Trials

ACT status cannot be determined directly from public ClinicalTrials.gov records because investigational new drug (IND) and investigational device exemption (IDE) status are not publicly available. Applicability was therefore estimated using a published, independently validated ACT proxy. Trials met the proxy if they were interventional, phase 2 or later, included a United States site, and evaluated an FDA-regulated drug or device. Because FDA regulation fields are incompletely populated in older records, applicability was assigned using confidence tiers: trials with populated FDA regulation fields were classified as flag-confirmed, those without were classified as inferred if they evaluated a drug, biologic, or device, and all others were considered non-applicable. Flag-confirmed and inferred groups are reported separately.

### Statistical Analysis

Categorical data are reported as frequencies with percentages, and continuous variables are represented as medians with interquartile ranges (IQRs). Reporting proportions are presented with Wilson 95% confidence intervals. Time from primary completion to first results submission was estimated using Kaplan–Meier methods, censoring trials without reported results at the extraction date; cumulative incidence is reported at 1, 2, and 5 years. Associations with timely reporting were evaluated using multivariable logistic regression including industry sponsorship, phase 2 or later, and log₂-transformed enrollment, with cluster-robust standard errors by lead sponsor. Reporting was also summarized by primary completion year (cohorts of ≥20 mature trials) relative to the 2007 FDAAA and 2017 Final Rule. National reporting estimates from DeVito et al. served as descriptive reference values.^8^ No adjustment for multiple comparisons was performed. Additional statistical details are provided in Supplementary File 1.

### Reproducibility and Independent Verification

Data extraction and analysis were performed by a single investigator using prespecified rules applied to structured registry fields, without manual abstraction or subjective classification. A second investigator, blinded to code development, independently reproduced the complete workflow on separate hardware using the archived query and confirmed that the reporting denominator and primary estimates met prespecified reproducibility thresholds (2% for the denominator and 3 percentage points for proportions). All reported figures are based on the original frozen dataset.

### Software

All data extraction and statistical analyses were performed in Python 3.13.9 using pandas 2.3.3, NumPy 2.3.5, Matplotlib 3.10.6, and statsmodels 0.14.5. ClinicalTrials.gov records were retrieved through the version 2 API using Python’s standard library. The complete extraction workflow, analysis code, and sponsor classification dictionary are publicly available in the project’s GitHub repository.

### Data Availability

The frozen study dataset, source code, quality-control documentation, decision logs, provenance record (hash: a94cc38b2751), and independent verification materials have been deposited on the OSF. ClinicalTrials.gov records are publicly available, and the archived repository contains the code and documented query required to reproduce all analyses.

## Results

### Cohort and trial flow

The search retrieved 5,851 completed or terminated interventional trials indexed to hypertension, and the retrieved count reconciled against the total declared by the API. Of these, 5,261 (89.9%) were completed and 590 (10.1%) were terminated. A total of 5,552 (94.9%) carried a primary completion date of type ACTUAL, and 5,396 of these were at least 12 months mature as of the extraction date and constituted the reporting denominator. 299 records were excluded for lacking an ACTUAL primary completion date and 156 for not yet being due (Figure 1).

**Figure 1.**
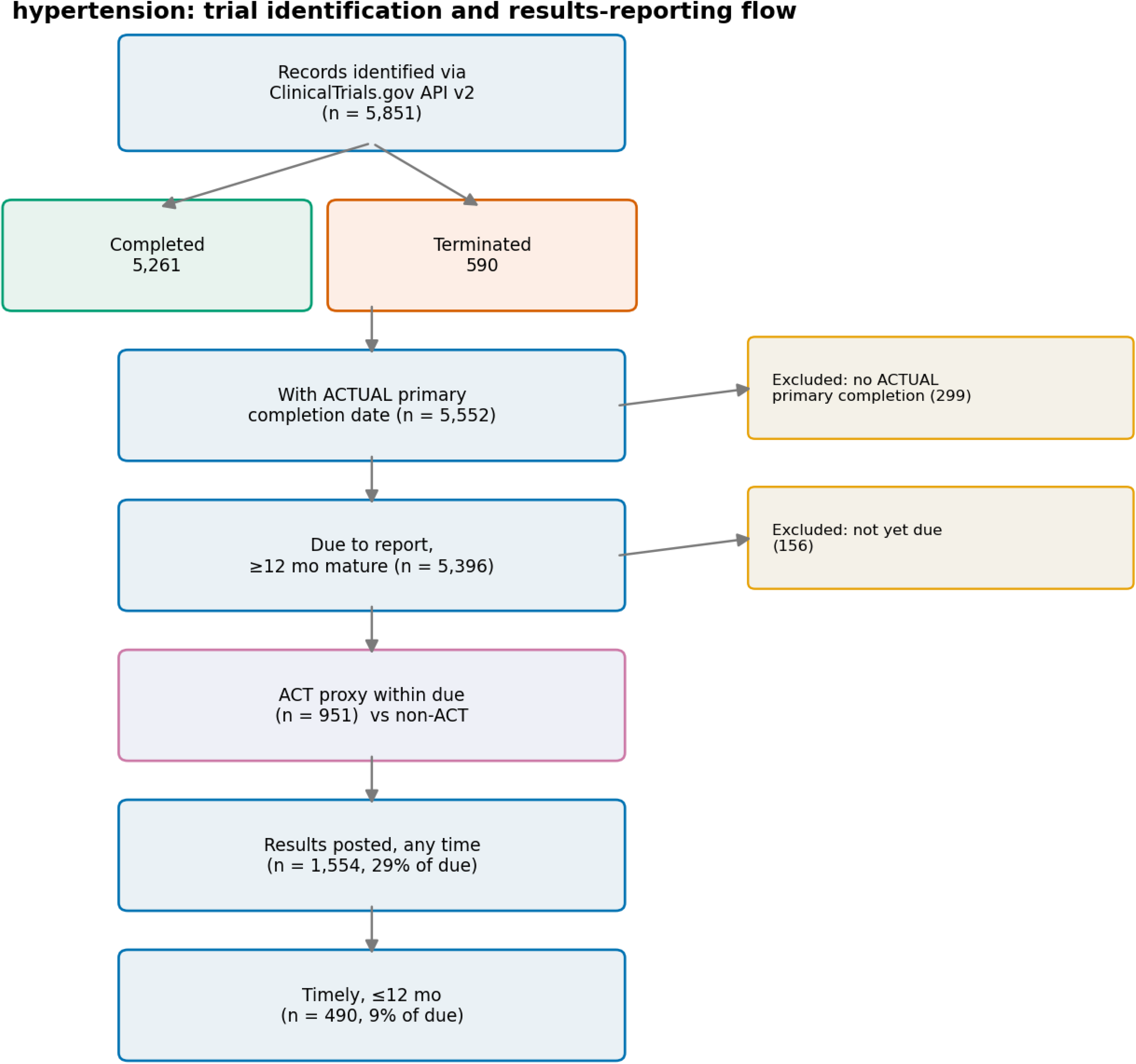
Identification and results-reporting flow for registered hypertension trials (PRISMA-adapted). The diagram shows retrieval from the ClinicalTrials.gov API, the completed and terminated split, records with an ACTUAL primary completion date, the due-to-report cohort (≥12 months mature), and the applicability, any-time, and timely reporting counts. The design uses a single registry source with no human screening stage; the figure is adapted from, and is not equivalent to, the PRISMA flow diagram.

### Trial characteristics

Among the 5,851 retrieved trials, 2,326 (39.8%) had at least one United States site and 2,730 (46.7%) were phase 2 or later. Median enrollment was 71 participants (IQR 31–202), and primary completion dates spanned 1979 to 2026. Lead sponsors were most often academic (2,205; 37.7%), industry (2,150; 36.7%), or other (466; 8.0%), with the remainder distributed across hospital (434; 7.4%), government (379; 6.5%), research non-profit (176; 3.0%), network (33; 0.6%), and individual (8; 0.1%) sponsors. Under the applicable-clinical-trial proxy, 322 trials (5.5%) were flag-confirmed and 702 (12.0%) were inferred, with the remaining 4,827 (82.5%) classified as non-applicable (Table 1).

**Table 1.** Characteristics of registered hypertension trials retrieved from ClinicalTrials.gov (N = 5,851).

| Characteristic | n | % or detail |
| --- | --- | --- |
| Completed/terminated interventional (total) | 5,851 | — |
| Completed | 5,261 | 89.9% |
| Terminated | 590 | 10.1% |
| With ACTUAL primary completion date | 5,552 | 94.9% |
| Due to report (ACTUAL, $\geq 12$ mo mature) | 5,396 | 92.2% |
| Has $\geq 1$ US site | 2,326 | 39.8% |
| Phase $\geq 2$ | 2,730 | 46.7% |
| ACT proxy — flag-confirmed | 322 | 5.5% |
| ACT proxy — inferred (flag-absent) | 702 | 12.0% |
| Non-ACT | 4,827 | 82.5% |
| Enrollment, median (IQR) | 71 | (31–202) |
| Primary completion year range | 1979–2026 | — |
| Sponsor: academic | 2,205 | 37.7% |
| Sponsor: industry | 2,150 | 36.7% |
| Sponsor: other | 466 | 8.0% |
| Sponsor: hospital | 434 | 7.4% |
| Sponsor: government | 379 | 6.5% |
| Sponsor: research non-profit | 176 | 3.0% |
| Sponsor: network | 33 | 0.6% |
| Sponsor: individual | 8 | 0.1% |

Values are n (% of total) unless otherwise noted. ACT, applicable clinical trial (proxy); IQR, interquartile range. The ACT proxy classifies all retrieved records; reporting-rate analyses use the due-to-report subset (Table 2a).

### Timely results reporting by applicability

Within the reporting denominator of 5,396 trials, 490 submitted summary results within 12 months of the primary completion date, a timely reporting rate of 9.1% (490/5,396; 95% CI 8.3– 9.9), below the Final-Rule reference of 40.9% (DeVito 2020). Trials meeting the applicable-clinical-trial proxy reported timely at 22.1% (210/951; 95% CI 19.6–24.8), while non-applicable trials reported timely at 6.3% (280/4,445; 95% CI 5.6–7.1), both below the reference. Within the applicable group, flag-confirmed trials reported timely at 36.9% (111/301; 95% CI 31.6–42.5), overlapping the reference when considering the confidence interval, whereas inferred trials, reported separately as lower-confidence, reported timely at 15.2% (99/650; 95% CI 12.7–18.2), below the reference (Figure 2, Table 2a).

**Figure 2.**
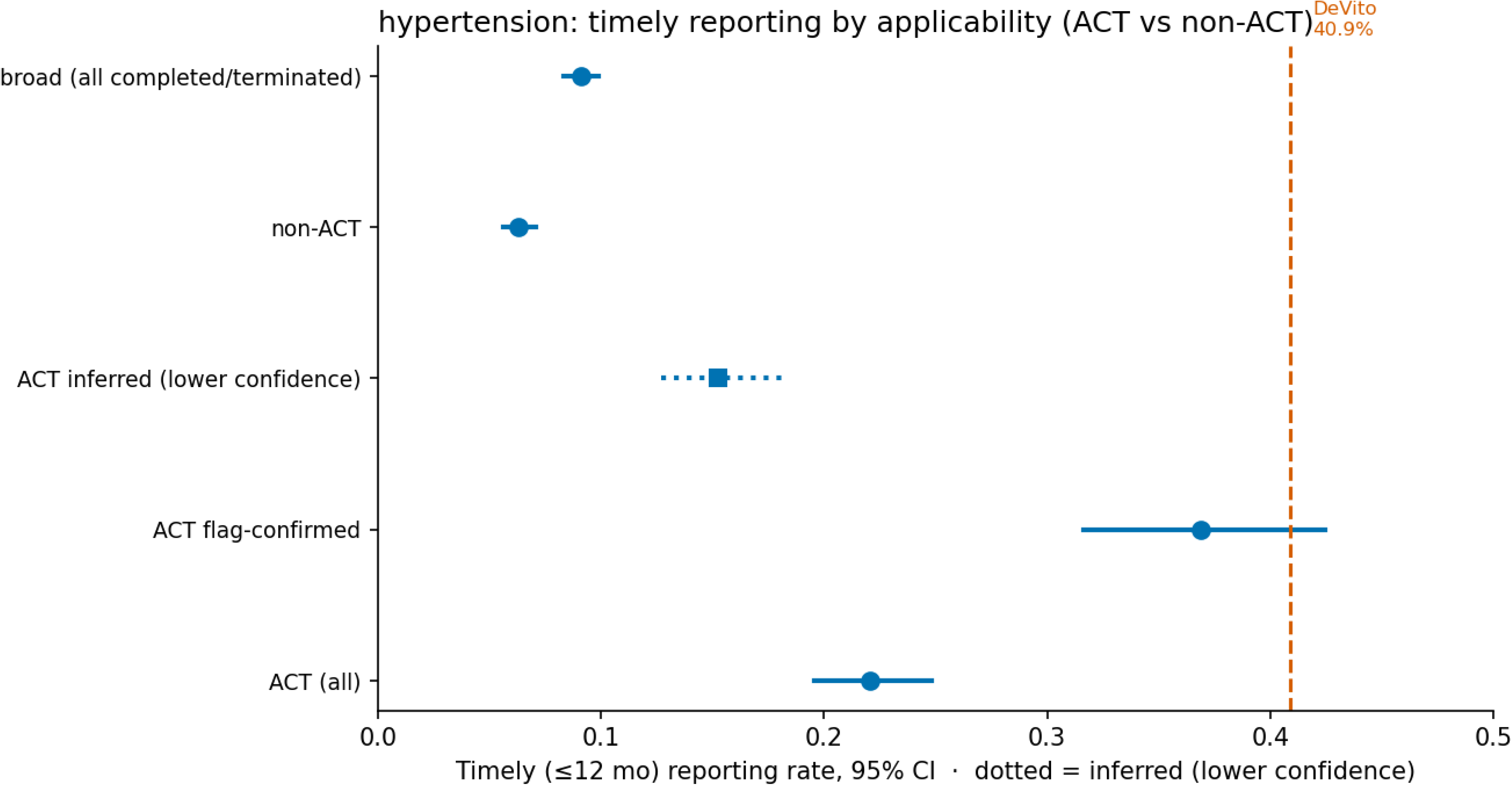
Timely (≤12 months) results reporting by applicability group, with 95% confidence intervals. The dashed line marks the Final-Rule national reference value (40.9%; DeVito 2020). Inferred applicable clinical trials are shown with a distinct marker and are reported as lower-confidence.

**Table 2a.**
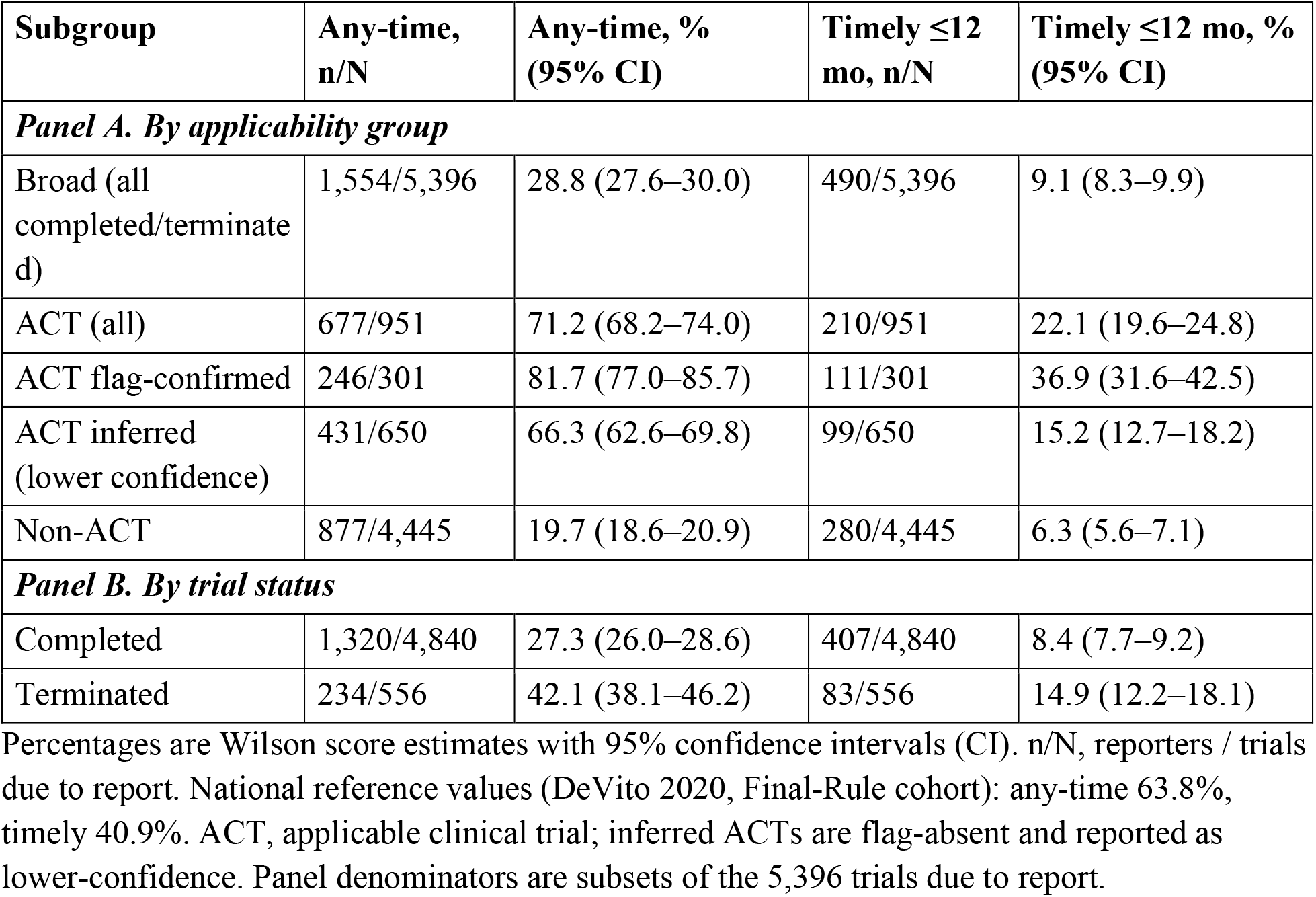
Results reporting, overall and by subgroup, among trials due to report (n = 5,396).

**Table 2b.** Adjusted associations with timely results reporting: multivariable logistic regression.

| Term | Adjusted OR (95% CI) |
| --- | --- |
| Industry sponsor | 1.72 (1.25–2.38) |
| Phase ≥2 | 1.86 (1.44–2.41) |
| Enrollment (per doubling) | 1.02 (0.96–1.09) |
| Has US site | 4.03 (2.99–5.42) |

n = 5,366 trials with complete covariates. OR, adjusted odds ratio with cluster-robust 95% confidence interval (1,673 lead-sponsor clusters). The model intercept is omitted.

### Any-time results reporting

Across the reporting denominator, 1,554 trials reported at any time, a rate of 28.8% (1,554/5,396; 95% CI 27.6–30.0), below the Final-Rule reference of 63.8% (DeVito 2020). Applicable trials reported at any time at 71.2% (677/951; 95% CI 68.2–74.0) and non-applicable trials at 19.7% (877/4,445; 95% CI 18.6–20.9). Within the applicable group, flag-confirmed trials reported at 81.7% (246/301; 95% CI 77.0–85.7) and inferred trials at 66.3% (431/650; 95% CI 62.6–69.8). The flag-confirmed and overall applicable groups exceeded the reference, the inferred group overlapped it, and the non-applicable group fell below it (Table 2a).

### Time to results reporting

Cumulative reporting incidence, estimated by the Kaplan–Meier method with administrative censoring at the extraction date, was 9.2% (95% CI 8.4–9.9) at 1 year, 19.0% (95% CI 17.9–20.0) at 2 years, and 27.3% (95% CI 26.1–28.5) at 5 years. Most trials never reported, so the survival function did not cross 0.5, and the median time to reporting was not reached (Supplementary Figure 1, Supplementary Table S1).

### Reporting by trial status

Completed trials reported at any time at 27.3% (1,320/4,840; 95% CI 26.0–28.6) and timely at 8.4% (407/4,840; 95% CI 7.7–9.2). Terminated trials reported at any time at 42.1% (234/556; 95% CI 38.1–46.2) and timely at 14.9% (83/556; 95% CI 12.2–18.1). Completed and terminated trials are reported separately (Table 2a).

### Sponsor type and adjusted associations

Industry-sponsored trials reported timely at 13.4% (268/2,001; 95% CI 12.0–15.0) and non-industry trials at 6.5% (222/3,395; 95% CI 5.8–7.4). In a multivariable logistic regression of timely reporting (n = 5,366), with cluster-robust standard errors by lead sponsor, the adjusted odds ratio was 1.72 (95% CI 1.25–2.38) for industry sponsorship, 1.86 (95% CI 1.44–2.41) for phase 2 or later, 1.02 (95% CI 0.96-1.09) per doubling of enrollment, and 4.03 (95% CI 2.99– 5.42) for the presence of a United States site (Table 2b).

### Reporting by completion-year cohort

Reporting was described by primary-completion-year cohort for cohorts of at least 20 mature trials, spanning 2003 to 2025, relative to the FDAAA 2007 and 2017 Final-Rule effective dates. Timely reporting rose across the period, from 0.0% in the earliest cohort (2003) to 16.6% in the 2024 cohort, whereas any-time reporting rose to a peak in the 2009 to 2013 cohorts and was lower in the most recent cohorts (Figure 3, Table 4). Because more recent cohorts have had less time to accrue reports, the any-time measure is interpreted with a follow-up caveat. The timely measure applies an identical 12-month window to every cohort and is the comparison that is fair across years.

**Figure 3.**
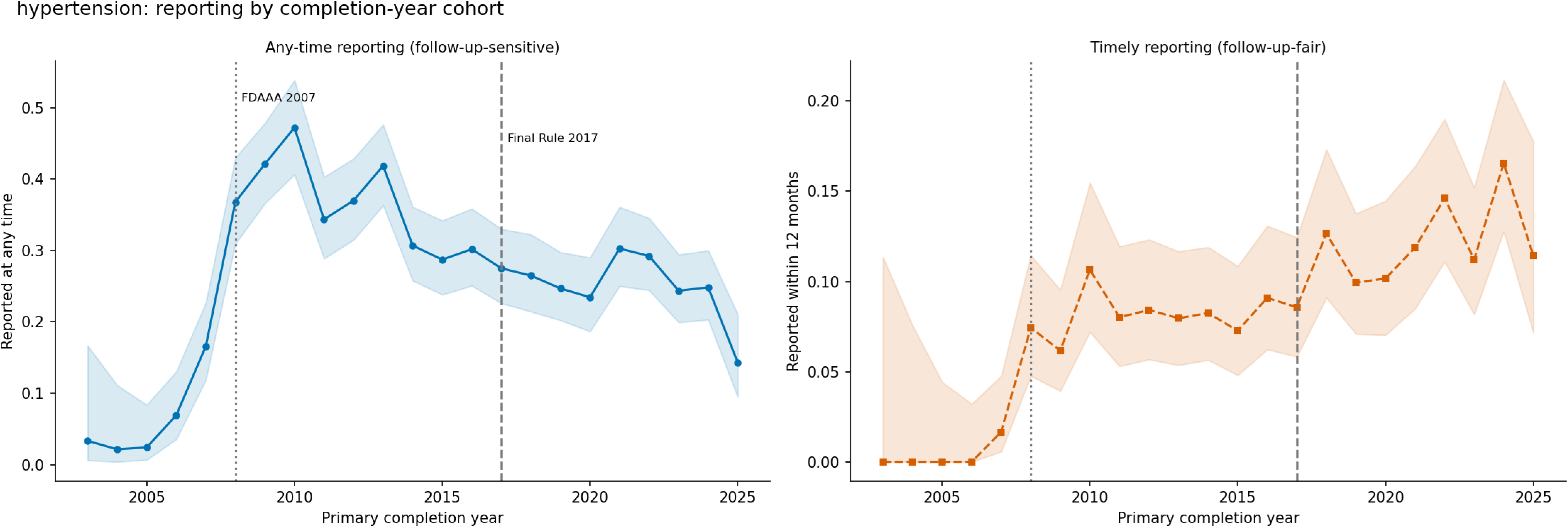
Results reporting by primary-completion-year cohort for any-time and timely (≤12 months) submission, restricted to cohorts of at least 20 mature trials and referenced to the FDAAA (2007) and Final-Rule (2017) effective dates.

### Submission-versus-posting sensitivity

The primary analysis timed submission of results, which is the action FDAAA obliges. Posting to the registry follows a National Library of Medicine quality-control review outside the responsible party’s control. Every reporter carried a results-first-submitted date, and no record required substitution of the posting date. Timing posting rather than submission lowered the broad timely rate roughly threefold, from 9.1% (490/5,396) to 3.0% (162/5,396; 95% CI 2.6– 3.5). The full comparison is given in Supplementary Table S2.

### Results in the literature but not the registry

Among the 3,842 trials due-to-report that had no posted results, a linked publication depended on how a link was defined. Under the strict definition, counting only sponsor-tagged results references, 7.8% (299/3,842; 95% CI 7.0–8.7) carried a linked publication. Under the broad definition, additionally counting auto-derived PubMed links, 36.4% (1,399/3,842; 95% CI 34.9– 38.0) did.

## Discussion

### Principal Findings

Timely reporting among registered hypertension trials was uncommon as less than 1 in 10 trials submitted results within the 12-month window, far below the national reference value.^8^ Restricting to trials meeting the ACT proxy raised these rates but did not close the gap. Taken together, these findings suggest that reporting behavior was more closely associated with whether a trial had a clearly identifiable reporting obligation than with characteristics of the condition itself. Hypertension is one of the most common chronic conditions worldwide, and incomplete or delayed reporting leaves the clinicians who treat it working from an incomplete evidence base.^1,15^ Trial results contribute to the evidence base only once publicly accessible, yet many here reported late or not at all.^16^

### The Applicability Gradient

The clearest divide in the data was between trials that carried a reporting obligation and those that did not. Because hypertension treatment relies predominantly on long-established therapies, relatively few registered trials evaluate novel FDA-regulated products under an active IND.^17^ Instead, many compare established therapies, test combinations or new populations, or evaluate lifestyle and device interventions. A large share are also academic or conducted outside the United States, consistent with the fact that less than half of this cohort had a US site. These are the trials the ACT definition does not reach, which is why more than four in five trials were non-applicable, and why the pooled reporting rate reflects the distribution of reporting obligations within hypertension research rather than reporting practices alone.

A non-applicable trial is not subject to a reporting deadline or associated penalties, so the incentive that drives timely posting does not apply. When analyzed separately, flag-confirmed trials reported at or above published benchmarks, whereas non-applicable trials reported substantially less frequently. The adjusted model revealed trials conducted at a US site carried the strongest association with timely reporting, ahead of later phase and industry sponsorship, while enrollment size had no effect. This held even though industry sponsored more than a third of the cohort, nearly as many trials as academia, so sponsorship did not substitute for an obligation. National audits found the same drivers at the registry level, where regulatory reach, later phase, and industry oversight predicted reporting.^8,18^

### Timeliness of Reporting

Applicable trials tended to report at some point, but often well behind schedule. These trials reported above the national any-time benchmark, but many did so only after the mandated 12-month window had closed. Across the full cohort, reporting accrued slowly and a typical time to reporting could never be defined, because most trials never reported at all. The delay matters, because antihypertensive guidance is built from pooled analyses of many trials and updated only every few years.^19,20^ A result that lands long after a trial finishes can miss the review or guideline it would have informed. Late reporting and non-reporting represent distinct problems, and addressing one will not necessarily resolve the other.

### Reporting Over Time

Timely reporting rose across successive completion-year cohorts, matching earlier national analyses.^21^ Any-time reporting looked highest in the middle of the period and lower in the most recent cohorts. That dip is an artifact of follow-up, because newer trials have had less time to report, and the fixed 12-month measure corrects it by giving every cohort the same window. The pooled estimate should be interpreted similarly. Since the cohort spans decades of hypertension research, the overall figure mixes eras with different reporting norms and understates where current practice stands.

### Measurement Considerations

Terminated trials were reported more often than completed ones, despite the common assumption that stopping early makes a trial more likely to go unreported. The obligation to report does not disappear when a trial stops early, and terminated trials may draw closer attention at closeout, though era and sponsorship differences could also contribute. Timely reporting also depended on which date was treated as the deadline. A sponsor submits results to the registry on one date, and those results become publicly visible on a later date after a quality-control review. Measuring from the submission date, the action the rule actually requires, produced a far higher timely rate than measuring from the later posting date. Additionally, a meaningful share of trials with no registry results had been published elsewhere, so registry-based counts may understate how often hypertension findings reach the literature. Nonetheless, result dissemination through publication fails to provide the raw evidence that the rule specifies and that guideline panels depend on.

### Implications

A single reporting rate for hypertension cannot be read as adherence because it pools obligated trials with a larger number that were not obligated. Separating them allows for distinct interpretation, which matters most for the people who use the registry to assemble the antihypertensive evidence base. Their search is weighted toward trials that carry an obligation and misses precisely what is often most decision-relevant in hypertension — the investigator-initiated comparison of established agents, the pragmatic target trial, and the head-to-head that no company is required to post. Among obligated trials, timeliness appears to be the most tractable challenge, as most eventually report results. In contrast, the trials least likely to report are also those over which the current reporting framework has the least leverage. This study evaluated no intervention and named no responsible party; it locates where the gap sits so effort can be aimed at it.

### Strengths and Limitations

This study examined the full set of completed and terminated interventional trials the registry indexes to the condition. This permits a within-condition comparison between trials that appear to carry a reporting obligation and those that do not. Every variable was derived from structured registry fields by fixed rule, the dataset was frozen, and the analysis was independently re-executed in order to maintain reproducibility. Applicability was assigned in graded tiers reported separately, so the effect of the inference is visible. Reporting was timed from submission, with posting as a sensitivity analysis.

Regulatory applicability cannot be read directly from public registry fields, because it depends partly on unpublished information. It stands on a validated proxy for the observable elements of the definition rather than an adjudication, a constraint shared by earlier assessments. Requiring a United States site renders the applicable set conservative, since it omits trials conducted entirely abroad under a domestic obligation. The cohort reflects the condition as the registry indexes it and it covers a single condition from one registry, so the findings may not extend to other conditions or registries. Comparisons with national figures are descriptive, and registry-based ascertainment misses results appearing only in the literature, a limitation quantified by the publication linkage in the results.

Future exploration could resolve applicability more precisely by linking registry records to regulatory sources. Transparency audits of newly approved drugs have already done this using public FDA review documents.^22^ Extending the linkage to regulatory information that is not publicly posted would close the distance between obligated and non-obligated trials.

### Conclusion

Prompt registry reporting of hypertension trial results remains uncommon, and how likely a trial was to report depended mostly on whether it was required to do so. Clinical trials with the clearest obligation reported at rates approaching the benchmark (36.9% timely), and the remaining shortfall reflects timing rather than omission, since any time reporting reached 71.2%. Nearly two decades after the publication of FDAAA 2007, timely reporting among hypertension trials has improved but remains incomplete. Much of the remaining gap occurs among trials without an identifiable reporting obligation despite their potential contribution to the evidence base.

## Author Contributions

William Harris and Parker Bragg performed data screening and drafted the original manuscript. Tanner Livsey contributed to conceptualization, including protocol design, Open Science Framework (OSF) registration, and Institutional Review Board (IRB) submission, and built and applied the data extraction framework. Ryan Langerman, Noah Calvert, Makalie Lackey, Lauren Kocour, and Amy Nguyen contributed to the construction and administration of the data extraction framework, the interpretation of data, and interactive education on epistemic safety and manuscript drafting. Alicia Ito Ford provided supervision, education, and project administration. Matt Vassar conceived the original project idea and provided supervision and project administration. All authors meet International Committee of Medical Journal Editors (ICMJE) authorship criteria and agree to be accountable for all aspects of the work.

## Data Availability Statement

All data supporting this study are publicly available. The underlying trial records were obtained from ClinicalTrials.gov, a public registry. The frozen analytic dataset, the complete extraction and analysis code, the sponsor classification dictionary, the quality-control and decision logs, the provenance record (stamp_hash”: “a94cc38b2751”), and the independent-verification materials have been deposited on the Open Science Framework [https://osf.io/sfgea] and archived on GitHub (methodology-c, commit c81585d, release hypertension-frozen-v3). Because the registry accrues records continuously, a fresh extraction returns an updated cohort; all reported figures derive from the original frozen extraction, and the analysis can be regenerated exactly from the archived code and the recorded query.

## Data Availability

https://osf.io/sfgea/overview?view_only=a9466f90be07439782e3b8d17d6a66fd

---

**Supplementary Table S1.**
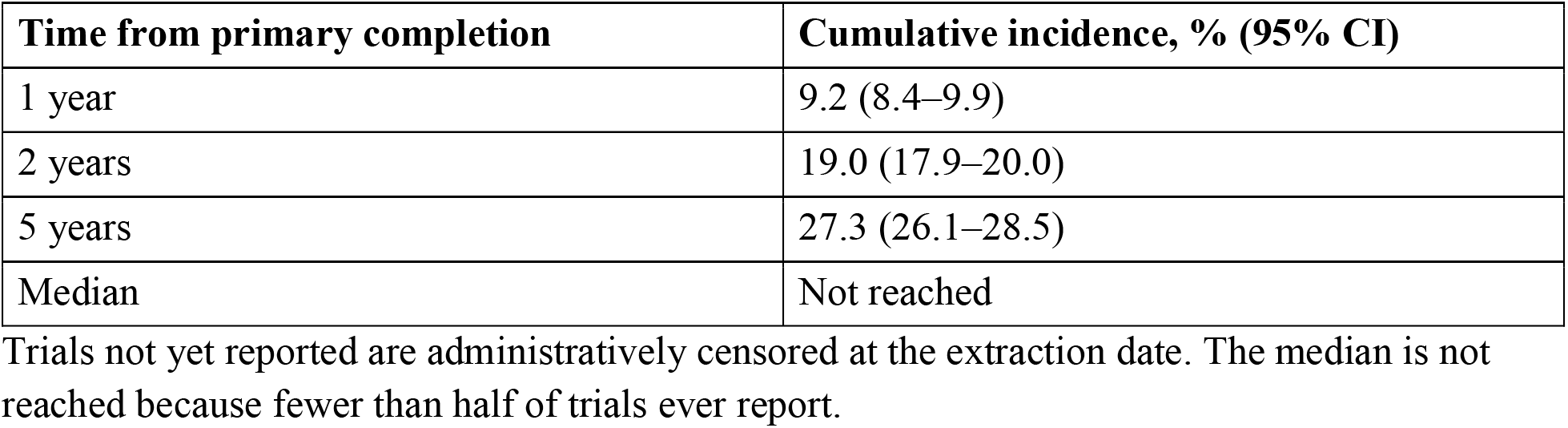
Kaplan–Meier cumulative results-reporting incidence.

| Time from primary completion | Cumulative incidence, % (95% CI) |
| --- | --- |
| 1 year | 9.2 (8.4–9.9) |
| 2 years | 19.0 (17.9–20.0) |
| 5 years | 27.3 (26.1–28.5) |
| Median | Not reached |

Trials not yet reported are administratively censored at the extraction date. The median is not reached because fewer than half of trials ever report.

**Supplementary Table S2.**
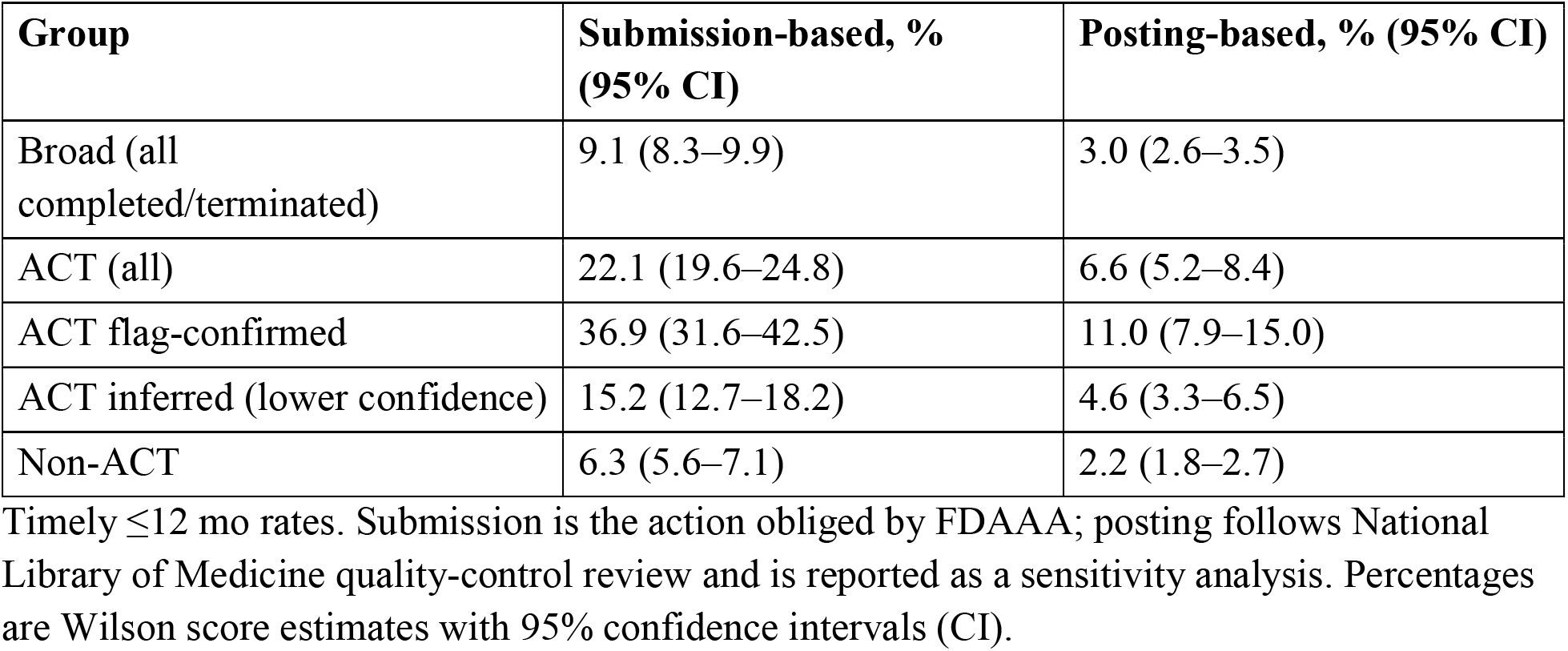
Timely results reporting under submission-based (primary) versus posting-based (sensitivity) clocks.

| Group | Submission-based, % (95% CI) | Posting-based, % (95% CI) |
| --- | --- | --- |
| Broad (all completed/terminated) | 9.1 (8.3–9.9) | 3.0 (2.6–3.5) |
| ACT (all) | 22.1 (19.6–24.8) | 6.6 (5.2–8.4) |
| ACT flag-confirmed | 36.9 (31.6–42.5) | 11.0 (7.9–15.0) |
| ACT inferred (lower confidence) | 15.2 (12.7–18.2) | 4.6 (3.3–6.5) |
| Non-ACT | 6.3 (5.6–7.1) | 2.2 (1.8–2.7) |

Timely $\leq 12$ mo rates. Submission is the action obliged by FDAAA; posting follows National Library of Medicine quality-control review and is reported as a sensitivity analysis. Percentages are Wilson score estimates with 95% confidence intervals (CI).

**Supplementary Table S3.**
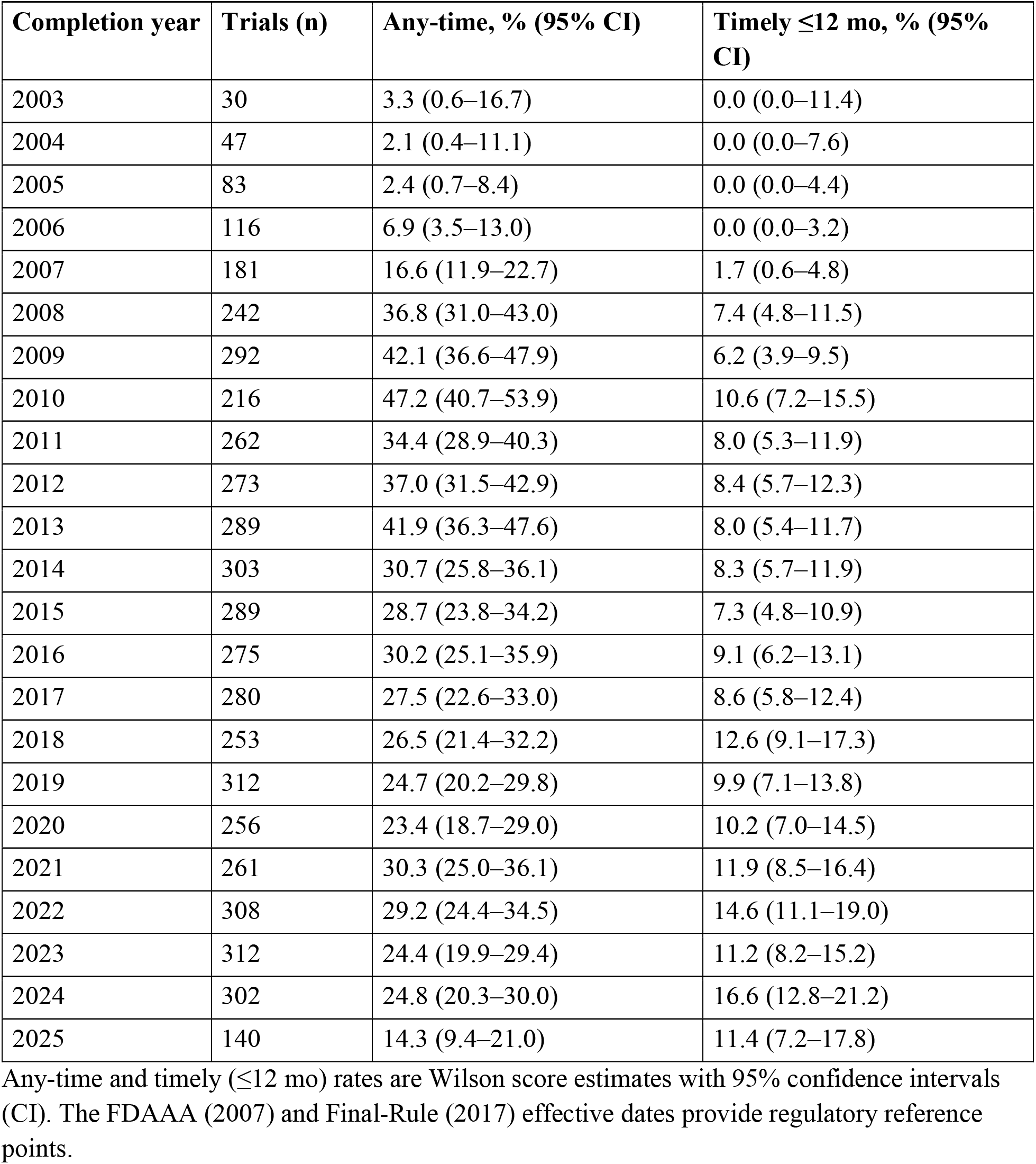
Results reporting by primary-completion-year cohort (cohorts of ≥20 mature trials).

**Supplementary Figure 1.** Kaplan–Meier cumulative incidence of results submission measured from the primary completion date, with trials not yet reported administratively censored at the extraction date. The 12-month regulatory deadline is indicated.

